# Reference values of the dorsal muscle group area for the diagnosis of sarcopenia using chest computed tomography in patients with chronic obstructive pulmonary disease: A prospective observational study

**DOI:** 10.1101/2025.09.21.25336291

**Authors:** Manabu Ono, Seiichi Kobayashi, Masatsugu Ishida, Hikari Sato, Koji Okutomo, Masaru Yanai

## Abstract

**Background:** Sarcopenia is common in patients with chronic obstructive pulmonary disease (COPD) and associated with poor prognosis. The dorsal muscle group area at the T12 vertebral level is useful in sarcopenia diagnosis; however, reference values for diagnosing sarcopenia are unavailable. This study aimed to establish the reference values for sarcopenia diagnosis using chest computed tomography scans in Japanese patients with chronic obstructive pulmonary disease.

**Methods:** We analyzed data from the Ishinomaki COPD Network Registry and selected the participants in the Ishinomaki COPD Network Registry who visited our hospital between April 2024 and March 2025. We investigated clinical parameters, including the dorsal muscle group area at the T12 vertebral level index (dorsal muscle group area at the T12 vertebral level/height^2^), to evaluate whether these parameters might serve as factors for sarcopenia.

**Results:** This study included 353 patients. According to the sarcopenia status, the patients were divided into the sarcopenia and non-sarcopenia groups. Propensity score matching and multivariate analyses were performed to compare baseline characteristics between the two groups. Multivariate regression logistic models revealed that the dorsal muscle group area at the T12 vertebral level index was an independent factor for predicting sarcopenia in patients with chronic obstructive pulmonary disease. The area under the receiver operating characteristic curve for identifying sarcopenia in patients with chronic obstructive pulmonary disease was 0.71, and the optimal cut-off value of the dorsal muscle group area at the T12 vertebral level index was 8.39.

**Conclusions:** Our study offers reference values for sarcopenia diagnosis using the dorsal muscle group area at the T12 vertebral level index evaluated using chest computed tomography in patients with chronic obstructive pulmonary disease. Identifying patients with chronic obstructive pulmonary disease in the early stages of sarcopenia can contribute to effective prevention and therapeutic measures.

## Introduction

Chronic obstructive pulmonary disease (COPD) is a chronic lung disease characterized by progressive airflow limitations and systemic inflammation and associated with multiple systemic manifestations that affect prognosis [1]. Patients with COPD exhibit alterations in body composition, muscle mass, and muscular function. Weight and muscle mass loss are associated with diminished muscle strength, exercise tolerance, and poor prognosis in patients with COPD [2,3]. Factors involved in the loss of muscle mass and function include oxidative stress, hypoxia, disuse, and malnutrition [4]. Reduced skeletal muscle mass (SMM) is associated with reduced pulmonary function [5,6]. Therefore, clinical assessment of SMM is important in patients with COPD. In elderly patients, COPD often presents with multimorbidities, including sarcopenia. Sarcopenia is defined as the age-associated loss of muscle mass and strength [7] and associated with poor quality of life and motility [8,9]. The prevalence of COPD-related sarcopenia is influenced by several risk factors, including the severity of lung disease and other clinical settings, such as systemic inflammatory response, oxidative stress, smoking, hypoxemia, long-term decreased activity, and malnutrition [10].

Diagnosis of sarcopenia is a complex process that requires specialized evaluation of three components: Muscle mass, strength, and physical function [7]. Although area measurement of muscle mass at the level of the third lumbar (L3) vertebra on a single cross-sectional computed tomography (CT) image is useful in evaluating sarcopenia [11], abdominal CT is not a routine examination for patients with COPD, and chest CT, as obtained in regular COPD assessments, does not reach beyond the first lumbar level (L1).

SMM analysis via chest CT demonstrates promising potential for the diagnosis of sarcopenia in patients with COPD because CT images acquired for other medical purposes can be repurposed to evaluate sarcopenia, or in patients who are unable to undergo bioelectrical impedance analysis to evaluate muscle mass. A decline in paravertebral muscle size and attenuation at the T12 vertebral level on CT images are associated with mortality among patients with hip fractures [12] and those undergoing liver transplantation [13]. A previous study reported reference values for sarcopenia diagnosis evaluated based on the dorsal muscle group (DMG) area at the T12 vertebral level (T12DMA) on CT images [14]; however, data on patients with COPD are scarce.

This study aimed to establish reference values for sarcopenia diagnosis through chest CT scans in Japanese patients with COPD. Our findings are anticipated to contribute to the development of effective preventive and therapeutic measures for sarcopenia-related medical problems.

## Materials and methods

### Study design

We conducted an observational cohort study to prospectively analyze data collected from consecutive scheduled visits or newly registered patients in the Ishinomaki COPD Network (ICON) Registry [15,16] between April 2012 and March 2025. Briefly, the ICON is a regional medical liaison system that aims to provide comprehensive care to patients with COPD and is part of a multicenter interdisciplinary collaboration among healthcare providers, specifically respiratory medicine specialists, nurse specialists, therapists, pharmacists, and general practitioners, in Ishinomaki, Japan. In accordance with COPD statements or guidelines [17,18], patients registered in the ICON Registry receive standard therapy and care in general practice clinics. Furthermore, patients undergo scheduled examinations and receive education at the Japanese Red Cross Ishinomaki Hospital every 6–12 months. Patients who experience exacerbations are first treated by their general practitioners and, if necessary, subsequently referred to the Japanese Red Cross Ishinomaki Hospital. The patient education program includes training in early exacerbation recognition and a written action plan for exacerbations using a self-management diary. Patients are prescribed short-acting bronchodilators, but not oral corticosteroids or antibiotics, for self-administration during exacerbations.

In this study, participants were selected from patients enrolled in the ICON Registry who visited our hospital between April 2024 and March 2025 (Fig 1).

**Fig 1.**
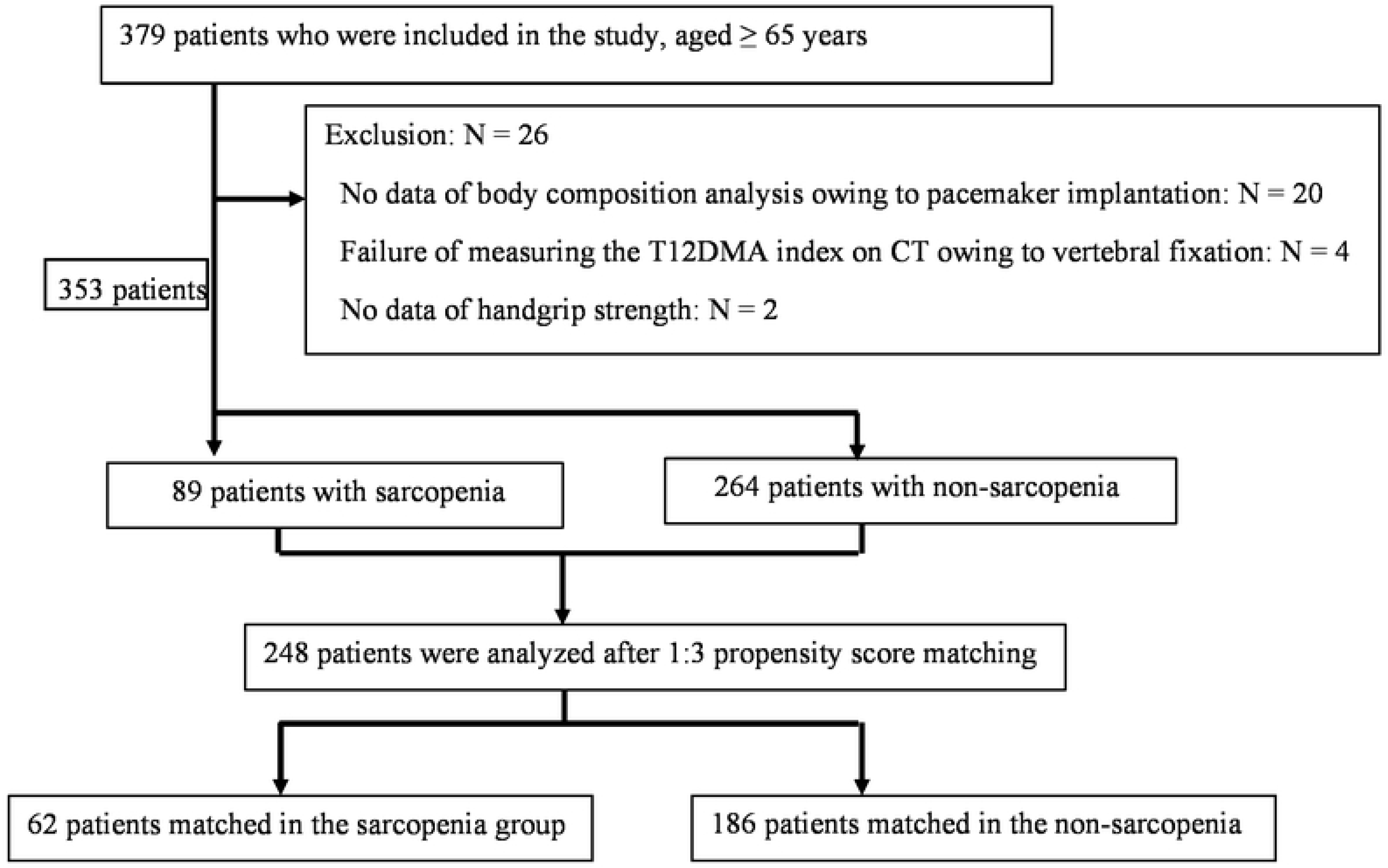
Flowchart of the Study. Patients with chronic obstructive pulmonary disease who were enrolled in the ICON and visited our hospital between April 2024 and March 2025 were selected. Abbreviation: ICON, Ishinomaki COPD Network; T12DMA, dorsal muscle group area at the T12 vertebral level; CT, computed tomography

### Ethical considerations

This study was part of an ongoing COPD cohort study registered with the University Hospital Information Network Clinical Trials Registry (identifier: UMIN000017376) and conducted in accordance with the 1964 Declaration of Helsinki and its later amendments or comparable ethical standards. All patients provided written informed consent, and this study was approved by the Ethics Committee of the Japanese Red Cross Ishinomaki Hospital (approval number: 12-14-1, 25-13).

### Patients

We enrolled patients with stable COPD (age range, 40–90 years) who were former smokers with a smoking history of at least ten pack-years at ICON. Patients with suspected COPD were referred to the Japanese Red Cross Ishinomaki Hospital. Patients diagnosed with COPD and confirmed to be in a stable condition were registered in the ICON registry if they provided consent. All patients were diagnosed with COPD based on the Global Initiative for Chronic Obstructive Lung Disease (GOLD) criteria. Repeated spirometry was used to confirm persistent airflow limitation, which is defined as a post-bronchodilator forced expiratory volume in 1 s (FEV1)/forced vital capacity ratio of < 0.7. The exclusion criteria were as follows: Current smoking, chronic bronchitis or emphysema without airflow limitation, use of oral corticosteroids or immunosuppressive or antifibrotic agents, hematological disease and COPD exacerbation within the 4 weeks preceding the data collection.

### Clinical and physiological measurements

We recorded each patient’s sociodemographic characteristics, smoking status, maintenance treatments, and history of malignant disease over the last 5 years. We also calculated the body mass index (BMI) in kg/m^2^ and evaluated dyspnea using the modified Medical Research Council dyspnea scale [17,18]. We assessed COPD-related health status using the COPD Assessment Test (CAT), an eight-item questionnaire with a possible total score of 0–40; a higher score indicates a worse quality of life [19,20]. The 6-min walk test (6MWT) was conducted according to the standardized protocol of the American Thoracic Society [21]. A well-trained technician conducted the pulmonary function tests following the standard guidelines under a stable condition [22]. We classified airflow limitation severity based on the GOLD staging as follows: GOLD I, FEV1 ≥ 80% predicted; GOLD II, 50% ≤ FEV1 < 80% predicted; GOLD III, 30% ≤ FEV1 < 50% predicted, or GOLD IV: FEV1 < 30% predicted [1]. We retrieved the levels of serum biomarkers, including albumin and total cholesterol, and radiological findings from the patients’ medical records.

Sarcopenia was diagnosed based on the loss of muscle mass, low muscle strength, and/or low physical performance according to the current criteria of the Asian Working Group for Sarcopenia (AWGS) 2019 [7]. Detailed methods and criteria are described below. Muscle mass: The SMM and SMM index (SMI) were calculated using a multifrequency bioelectrical impedance analyzer (InBody 770; InBody Japan, Tokyo, Japan). The cut-off values for low muscle mass were < 7.0 kg/m^2^ for men and < 5.7 kg/m^2^ for women. Muscle strength: Handgrip strength was measured in the standing position with full elbow extension using a Smedley-type hand dynamometer (Takei Scientific Instruments, Niigata, Japan). The measurements were obtained twice for each hand, with the largest value used as the grip strength value for analysis. The cut-off values for low muscle strength were < 28.0 kg for men and < 18.0 kg for women.

CT images were used to determine the quantity of skeletal muscle. The T12DMA was defined as the muscle within the region at the T12 vertebral level (Fig 2). Quantitative assessment of T12DMA was performed on a single axial slice of the CT image at the level of the 12th thoracic spine and measured with a manual sketch ridge of the DMG. The T12DMA was recorded as the sum of the bilateral DMG areas (Table 1). Because absolute muscle mass is strongly correlated with height, we evaluated the T12DMA index as an index of the relative SMM, which was calculated as T12DMA divided by the square of body height in meters.

**Fig 2.**
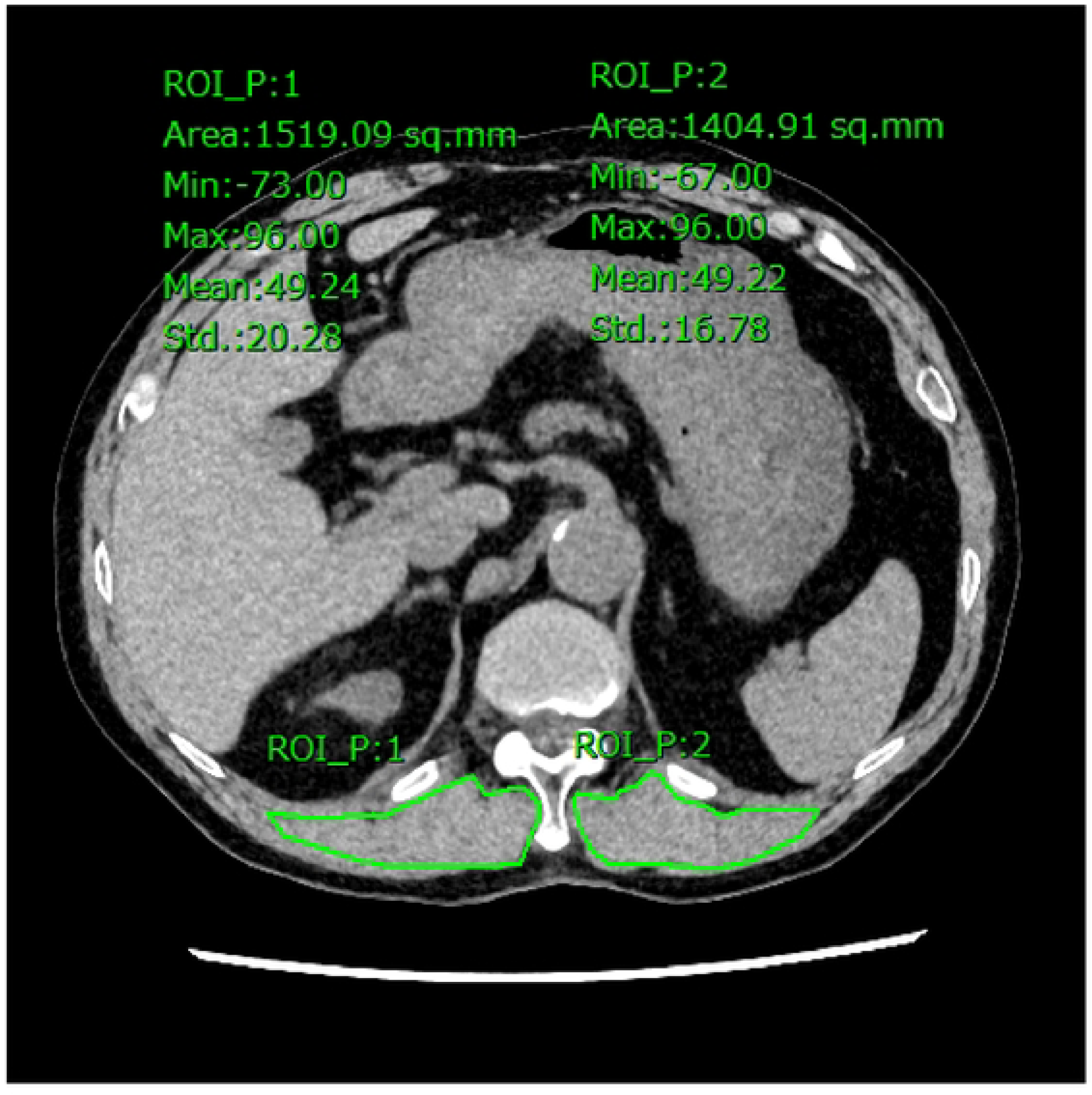
Representative Image of the T12DMA. The region marked with a solid line indicates the bilateral DMA. Abbreviation: T12DMA, dorsal muscle group area at the T12 vertebral level

**Table 1.**
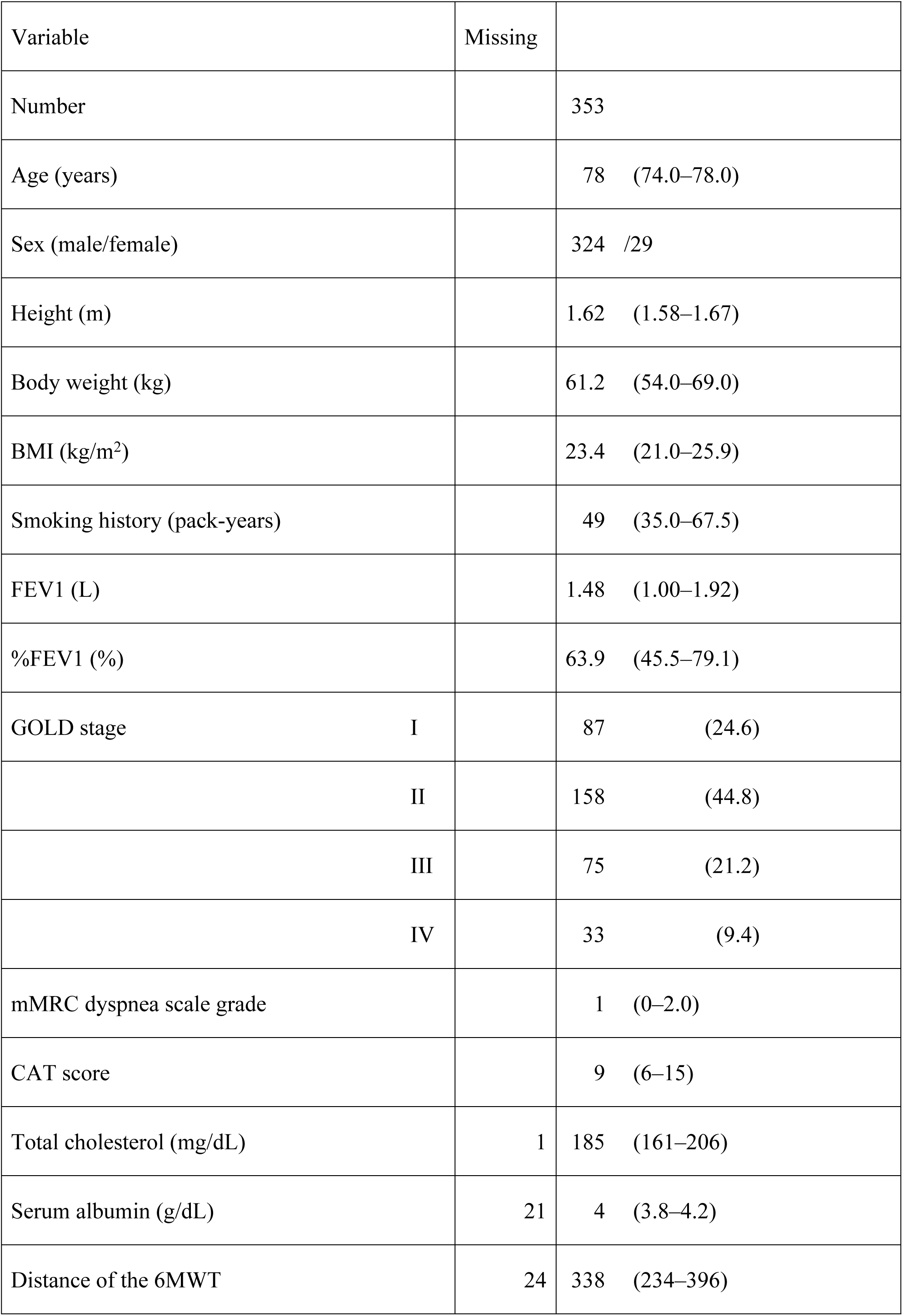
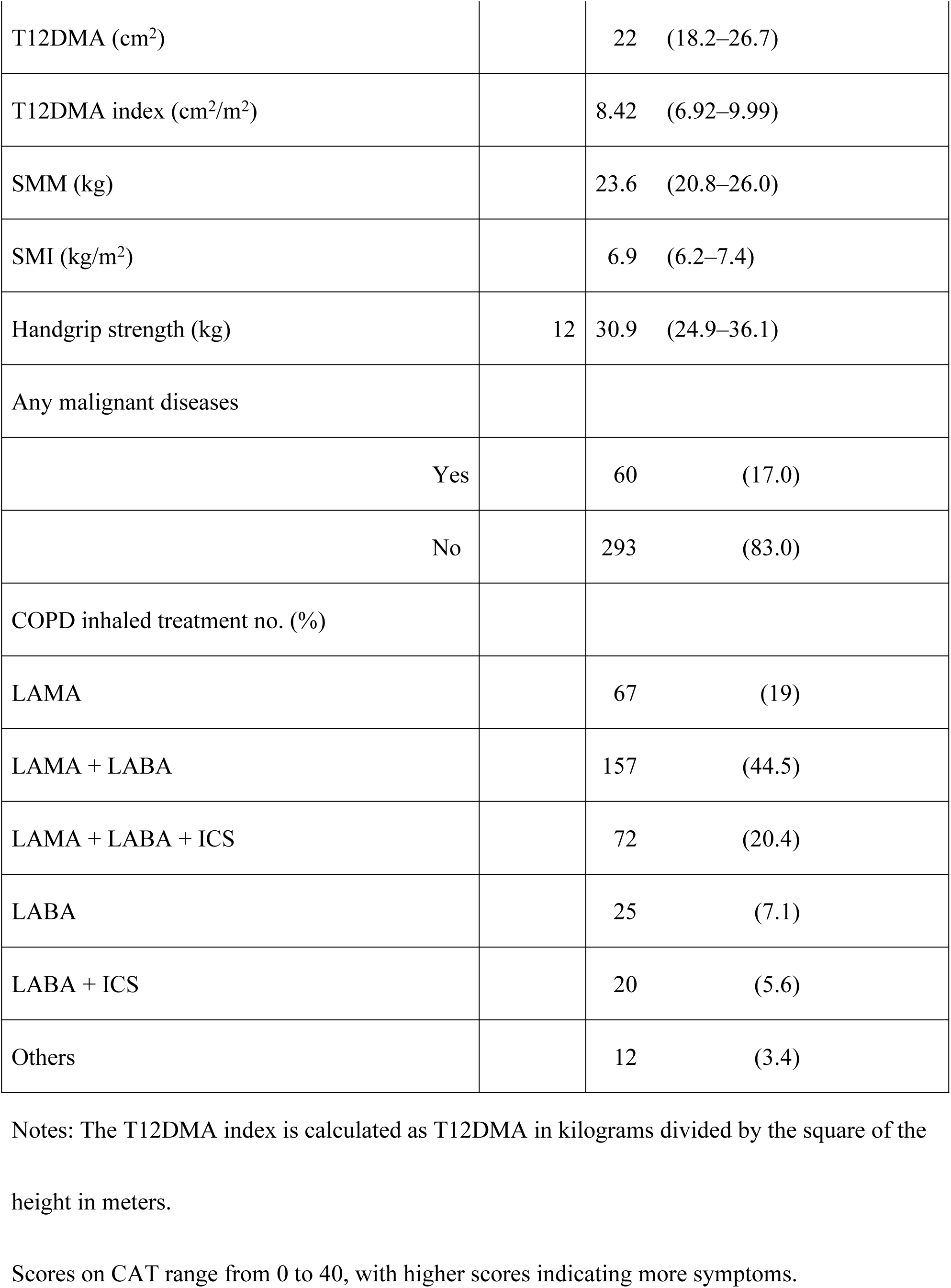
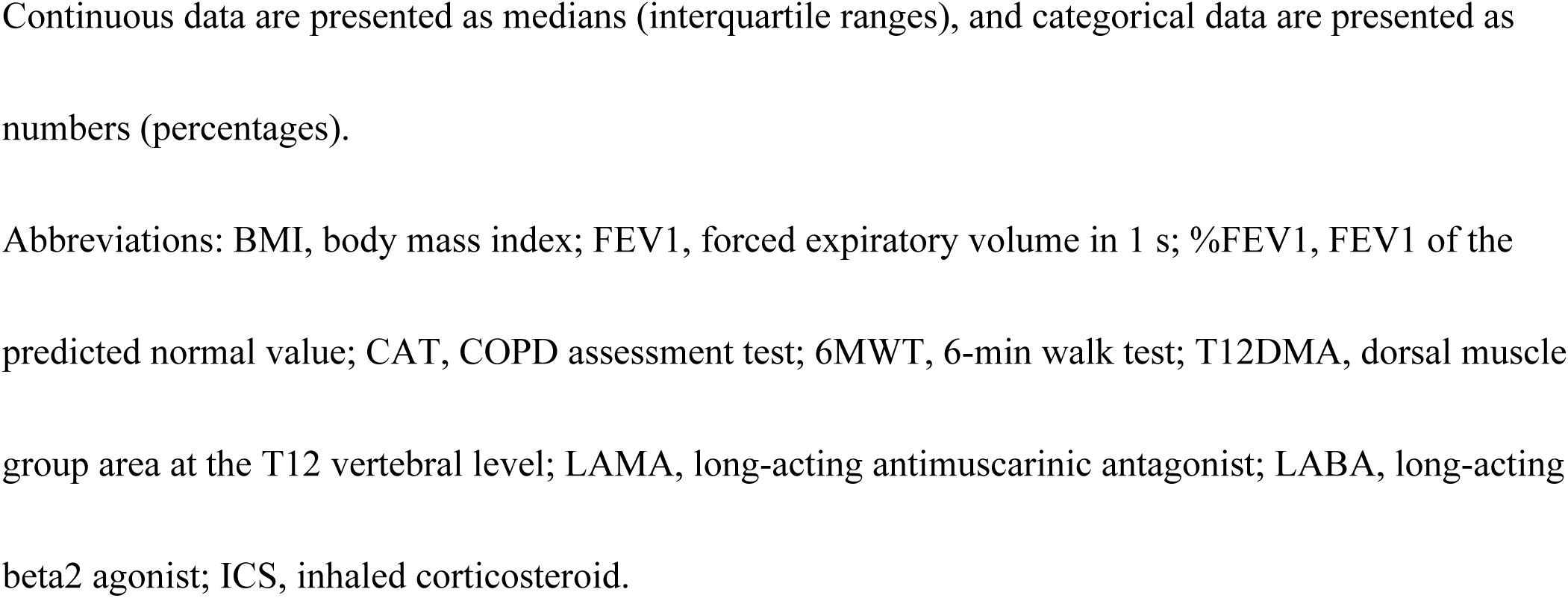
Patients’ Demographic and Clinical Data.

### Statistical analyses

The patients were categorized into two groups: Patients with COPD and sarcopenia (sarcopenia group) and patients with COPD without sarcopenia (non-sarcopenia group). Categorical data are presented as numbers (percentages) and were compared using Fisher’s exact test. Continuous data are presented as medians and 25– 75% interquartile ranges (IQRs) and were compared using the Mann–Whitney U test. The last observation was used to impute missing values. Continuous variables were analyzed using the Kruskal‒Wallis test with correction using the Steel test to compare differences among three or more groups. Propensity score matching (PSM) was implemented before statistical analysis to achieve balanced exposure at baseline for specific covariates that may affect the outcomes. The covariates included age, sex, and %FEV1 (Fig 1). The patients were 1:3 PS-matched using the near neighbor methodology with a maximum caliper of 0.02 of the deviation of the logit of the PS. Multivariate logistic regression analysis was performed after adjusting for factors, including variables found to be of interest in previous studies (particularly albumin level, BMI, T12DMA index). A receiver operating characteristic (ROC) curve was constructed and used to determine the cut-off values.

We performed all statistical analyses using EZR (Saitama Medical Center, Jichi Medical University, Saitama, Japan), which is a graphical user interface for R (the R Foundation for Statistical Computing, Vienna, Austria); P < 0.05 was considered significant [23].

## Results

In this study, patients with COPD who were enrolled in the ICON Registry and visited our hospital between April 2024 and March 2025 were selected according to the flowchart shown in Fig 1. A total of 379 patients aged ≥ 65 years were assessed for eligibility from April 2024 to March 2025 in our hospital; of these, 353 were identified as eligible for study inclusion. Twenty-six patients were excluded from this study for the following reasons: Body composition analysis was not performed in 20 patients owing to pacemaker implantation; the T12DMA on CT images of four patients could not be measured because the patients underwent vertebral fixation; and there was a lack of information for two patients. Demographic and baseline characteristics of the full cohort are presented in Table 1.

Briefly, the median age of the enrolled patients was 78.0 (IQR: 74.0–83.0) years. Men accounted for 91.8%, and the median BMI was 23.4 (IQR, 21.0–25.9) kg/m^2^. The median FEV1 was 1.48 L (IQR, 1.00–1.92), the median %FEV1 was 71.7% (IQR, 45.6– 79.1), and the serum albumin level was 3.8 (IQR, 3.5–4.0) g/dL. The median T12DMA was 22.0 (IQR, 18.2–26.7) cm^2^, and the T12DMA index was 8.41 (IQR, 6.92–9.99) cm^2^/m^2^.

Among patients with COPD, the T12DMA index decreased and correlated with disease severity (Fig 3). Patients with COPD with GOLD stages III (P = 0.02) and IV (P = 0.03) exhibited significantly lower values than patients with stage I COPD.

**Fig 3.**
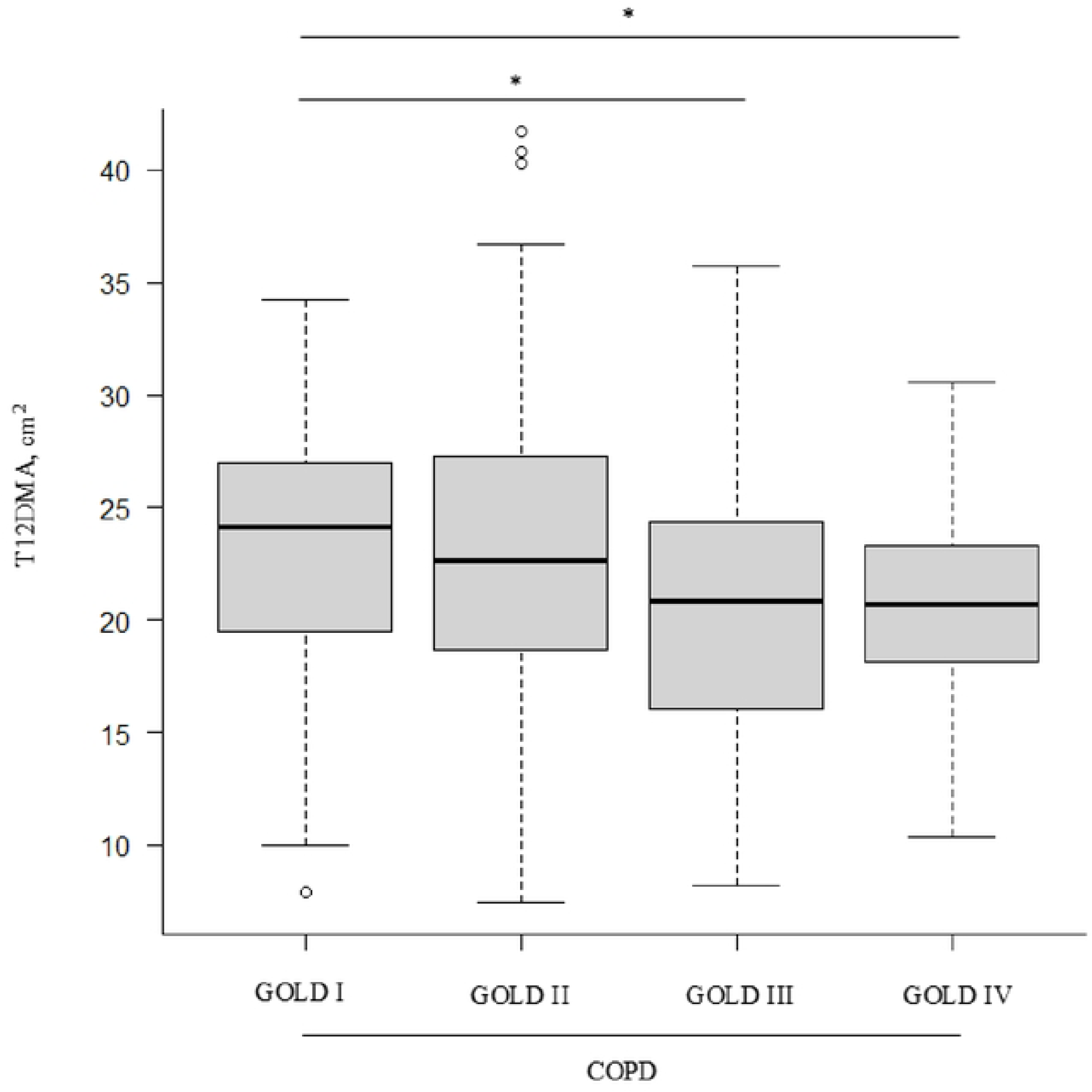
T12DMA Analyzed Using CT Images in Patients With COPD. The Kruskal-Wallis test with Steel’s post hoc test was used. *P < 0.05 compared with COPD GOLD. **I.** Abbreviations: T12DMA, dorsal muscle group area at the T12 vertebral level; CT, computed tomography; COPD, chronic obstructive pulmonary disease; GOLD, Global Initiative for Chronic Obstructive Lung Disease

**Fig 4.**
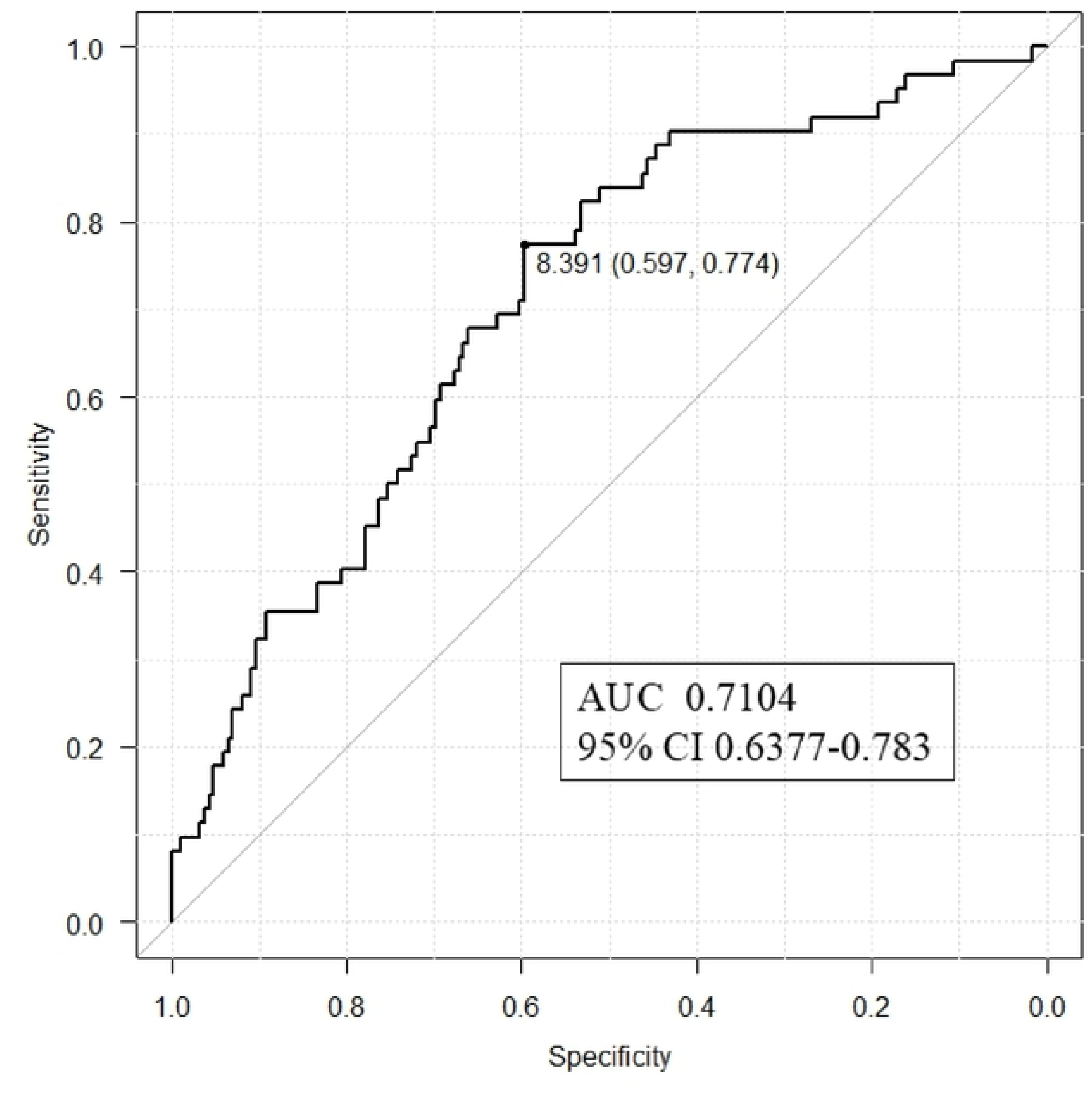
ROC of the T12DMA Index for Predicting Sarcopenia in Patients With COPD. Abbreviation: ROC, receiver operating characteristic curve; AUC, area under the curve; T12DMA, dorsal muscle group area at the T12 vertebral level.

Based on the diagnosis of sarcopenia, the patients were divided into two groups: Patients with COPD and sarcopenia, classified as the sarcopenia group (n = 89), and patients with COPD without sarcopenia, classified as the non-sarcopenia group (n = 264). The data were compared between the two groups. The comparisons between the sarcopenia and non-sarcopenia groups are shown in Table 2.

**Table 2.**
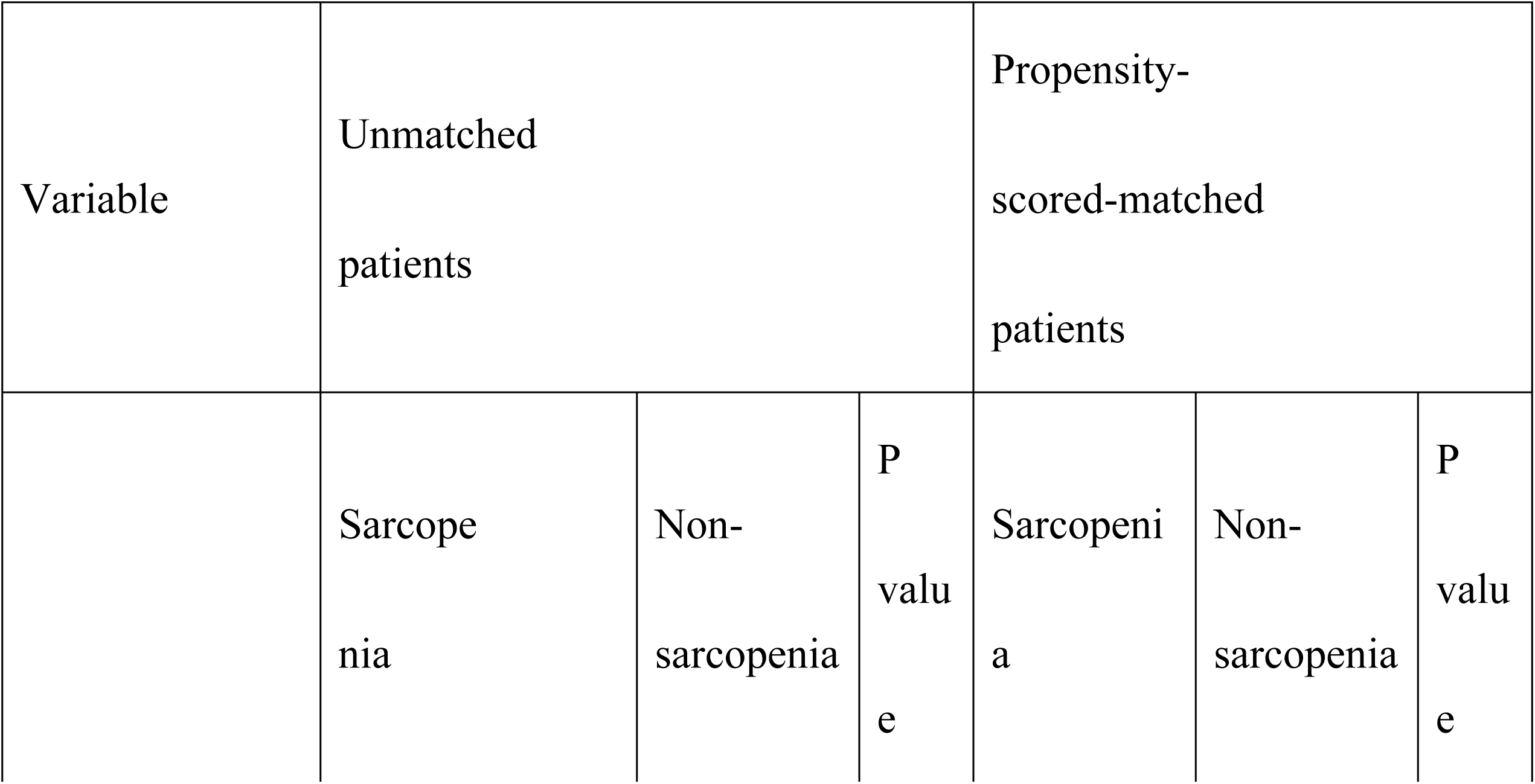
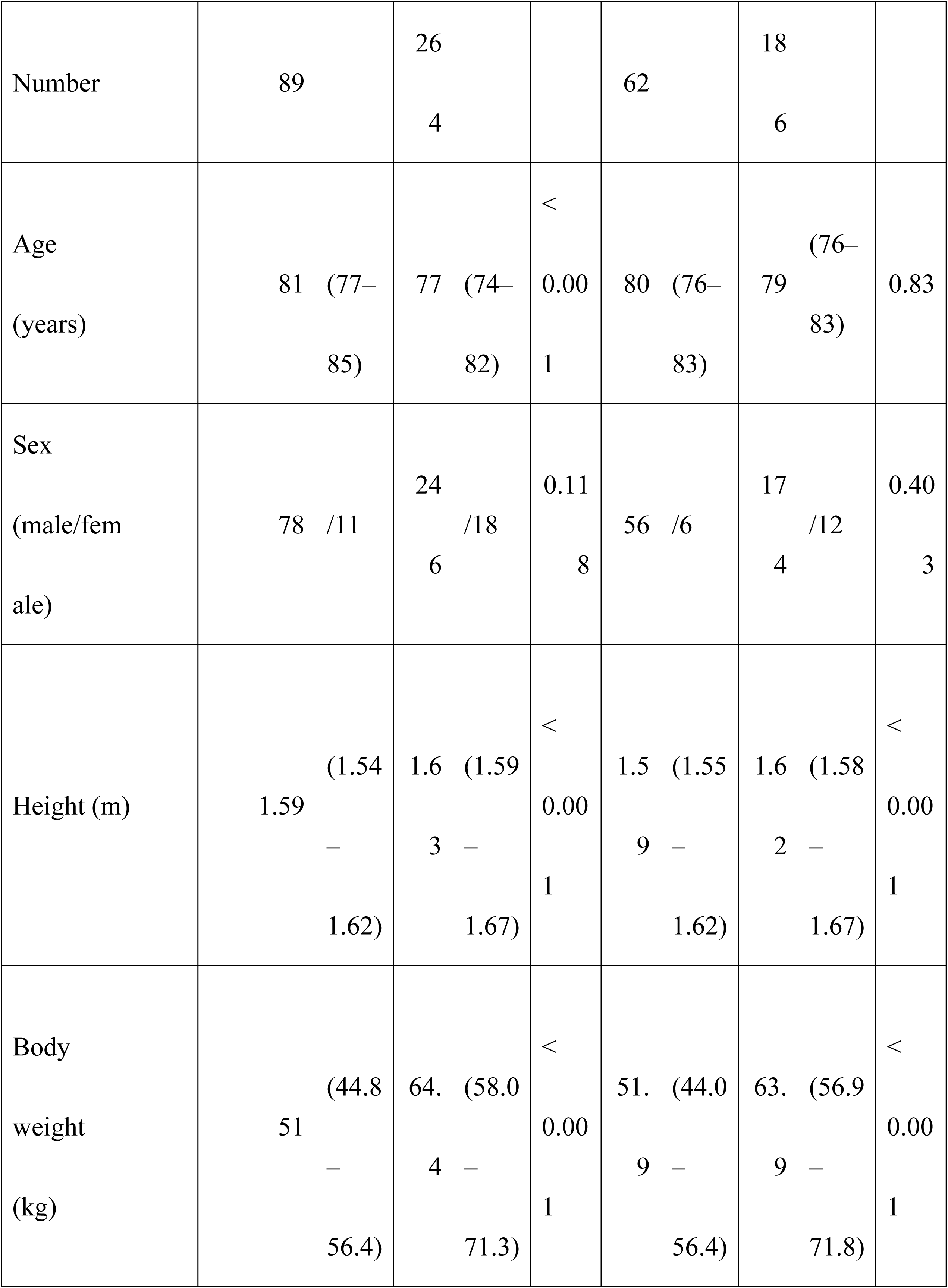
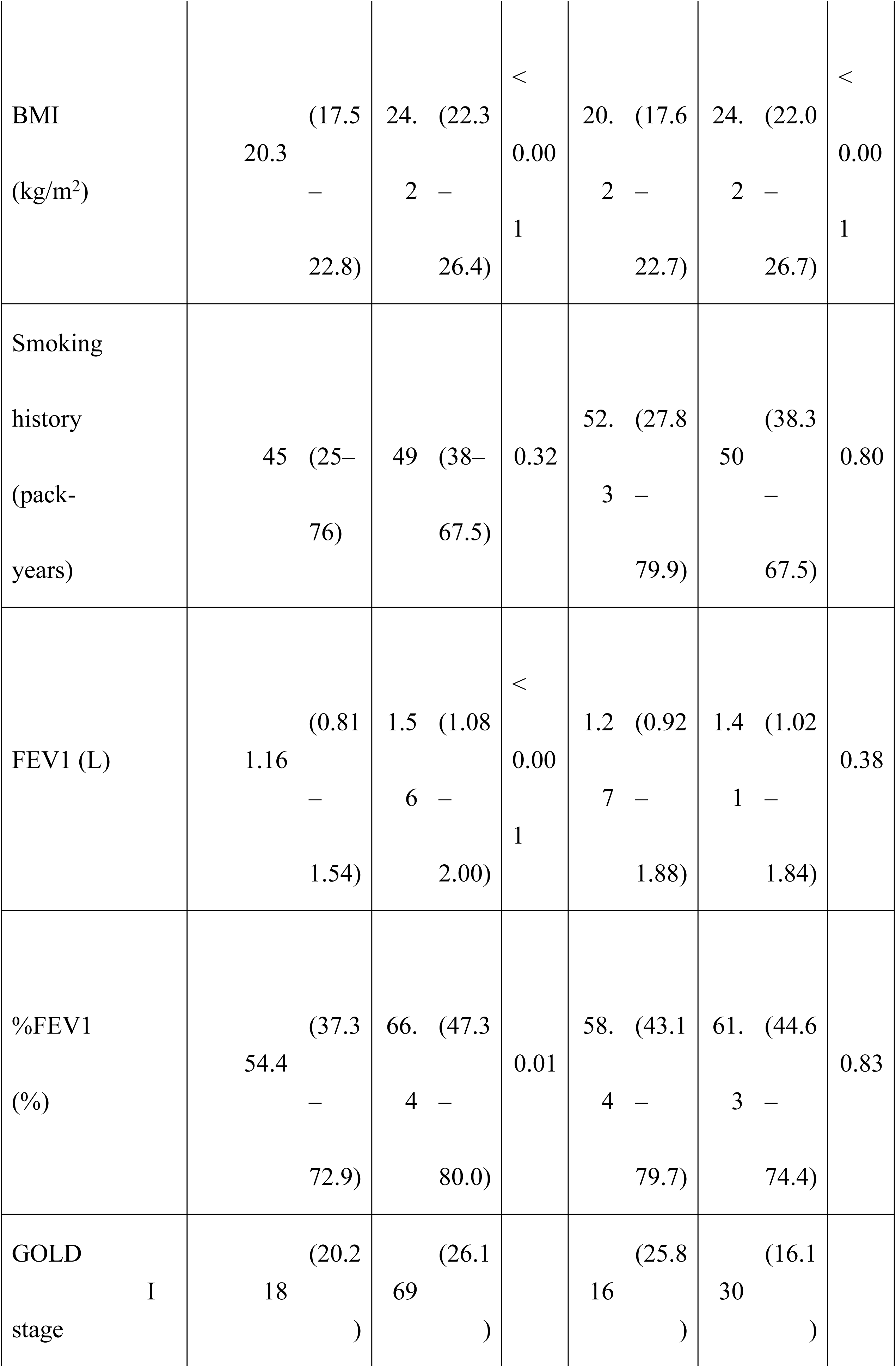
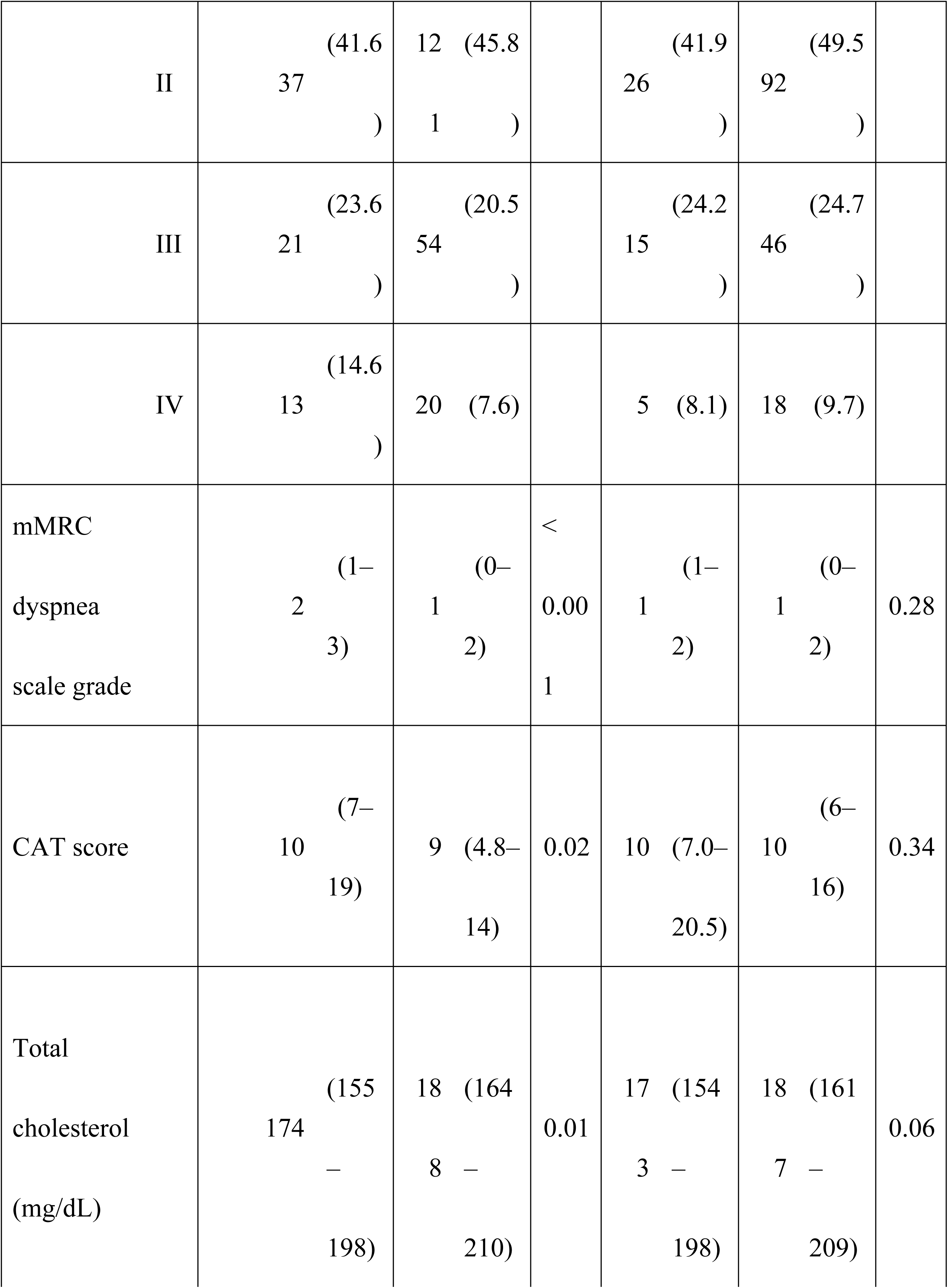
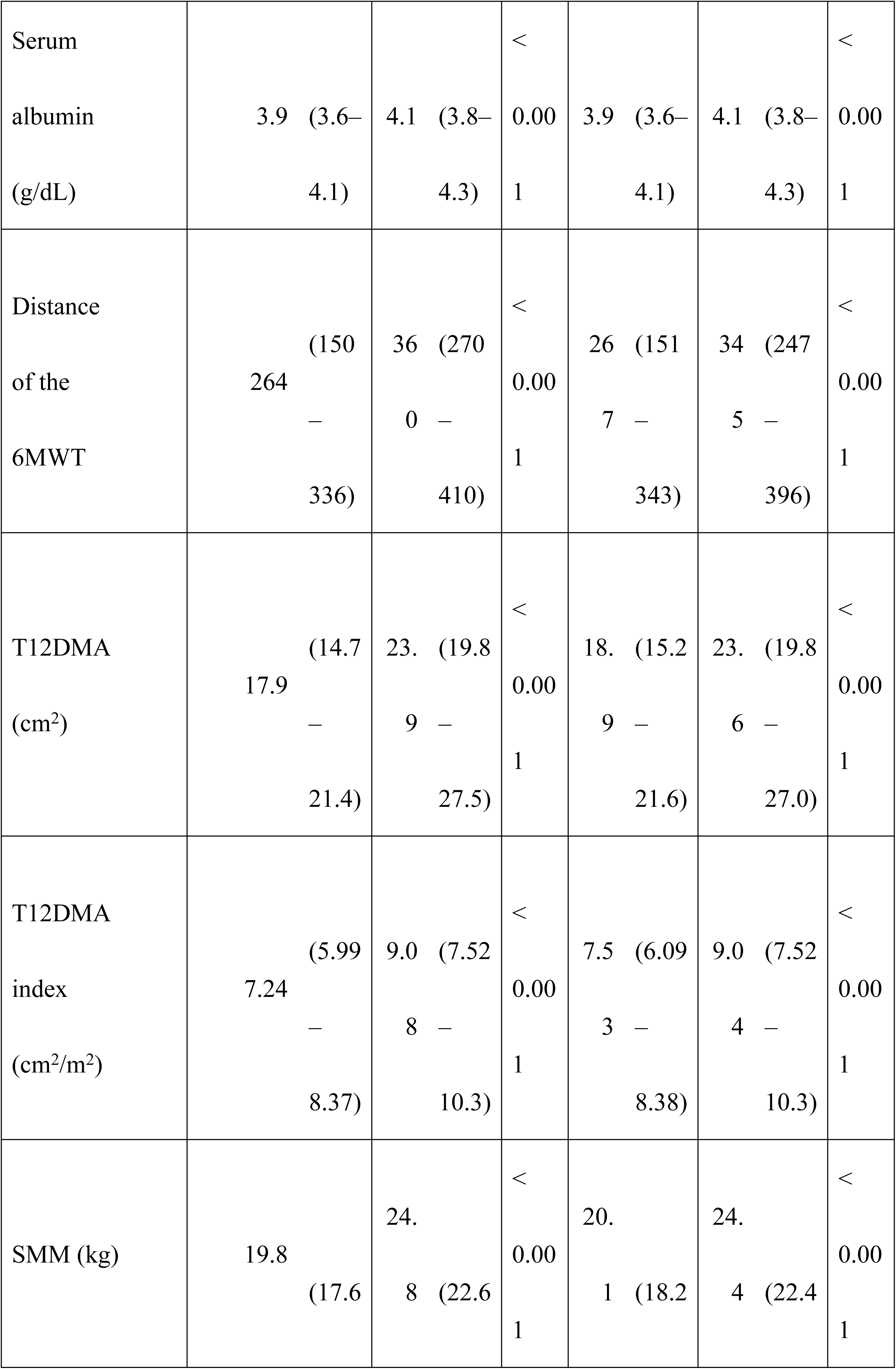
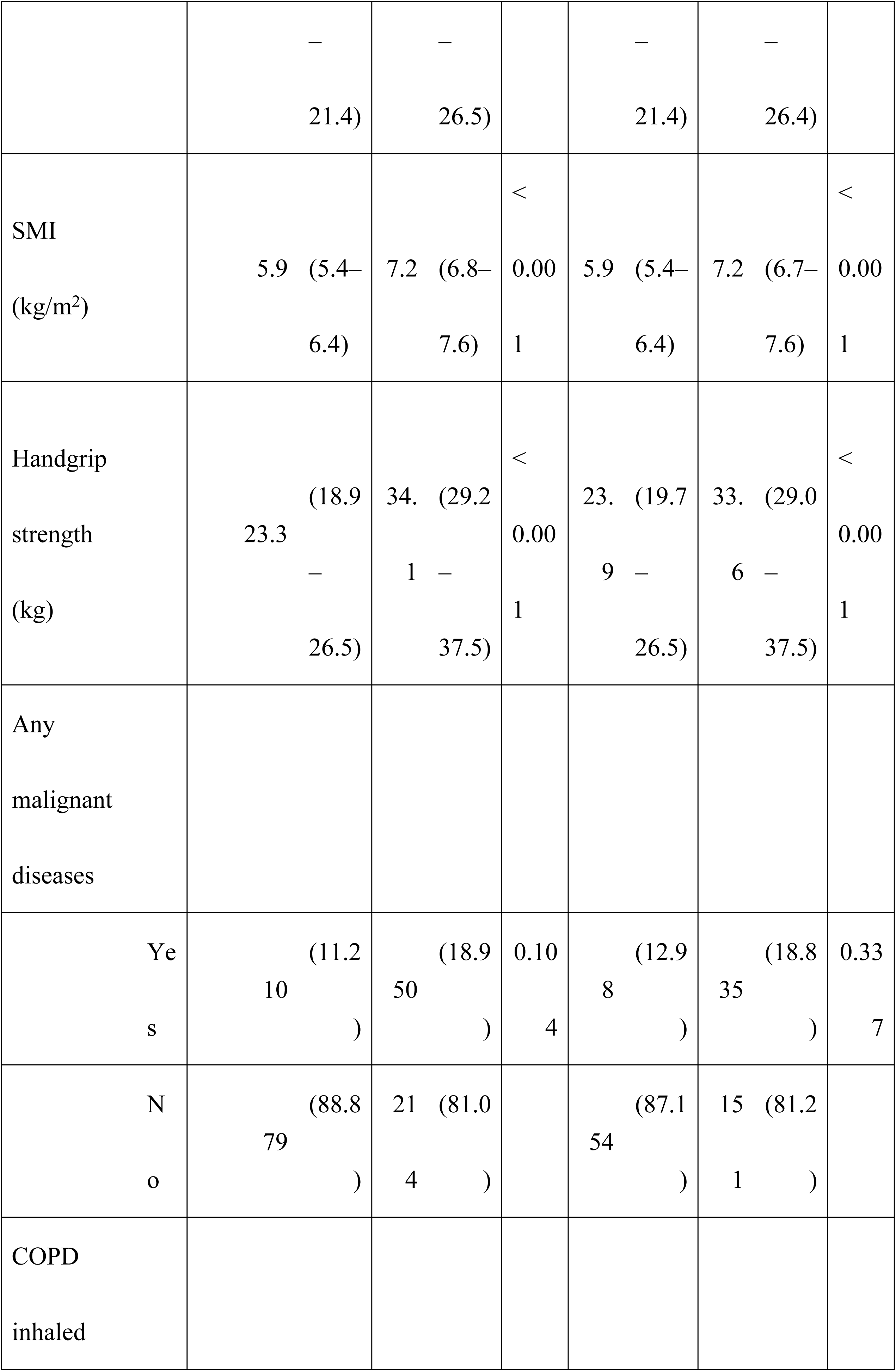
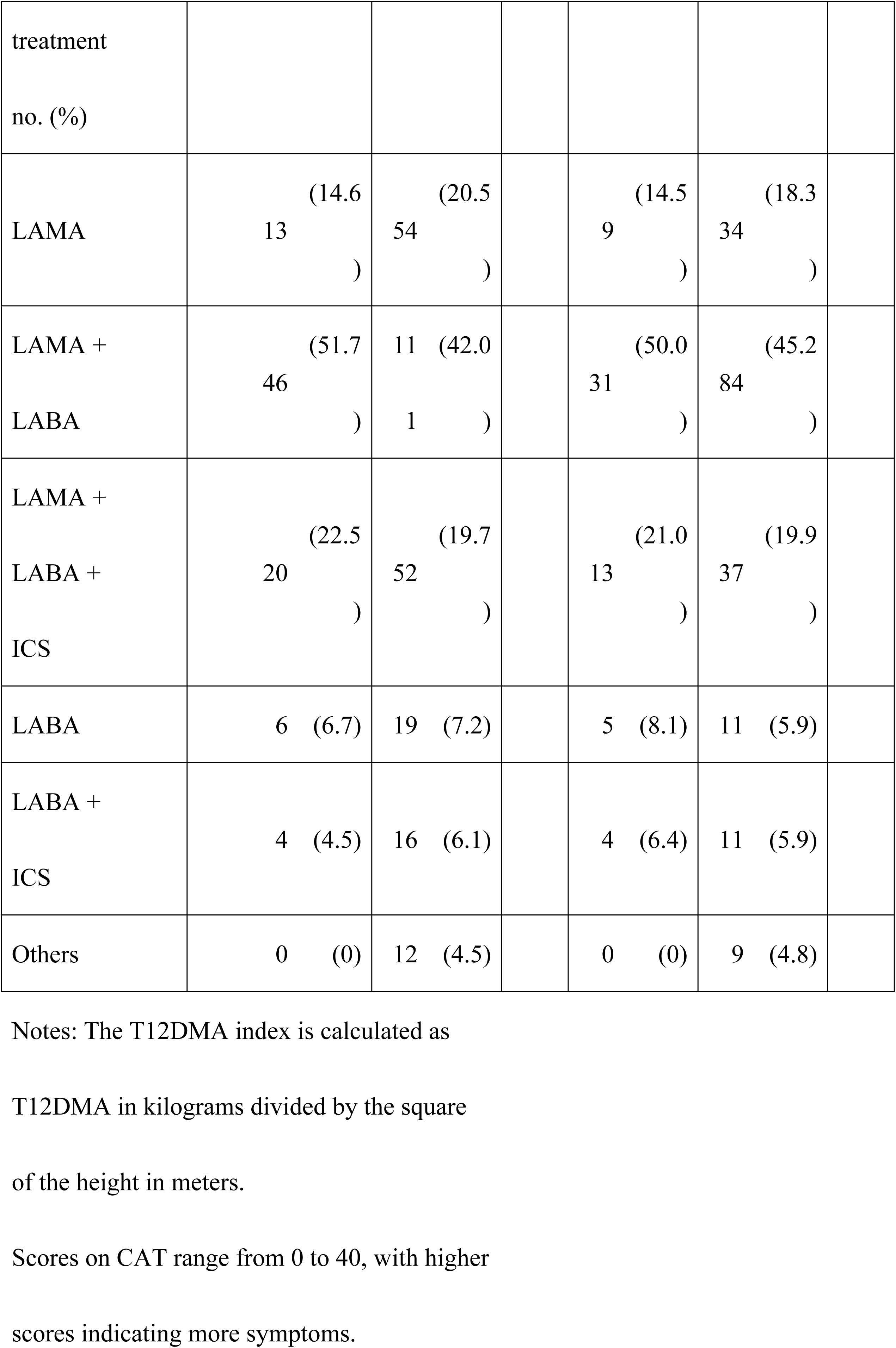
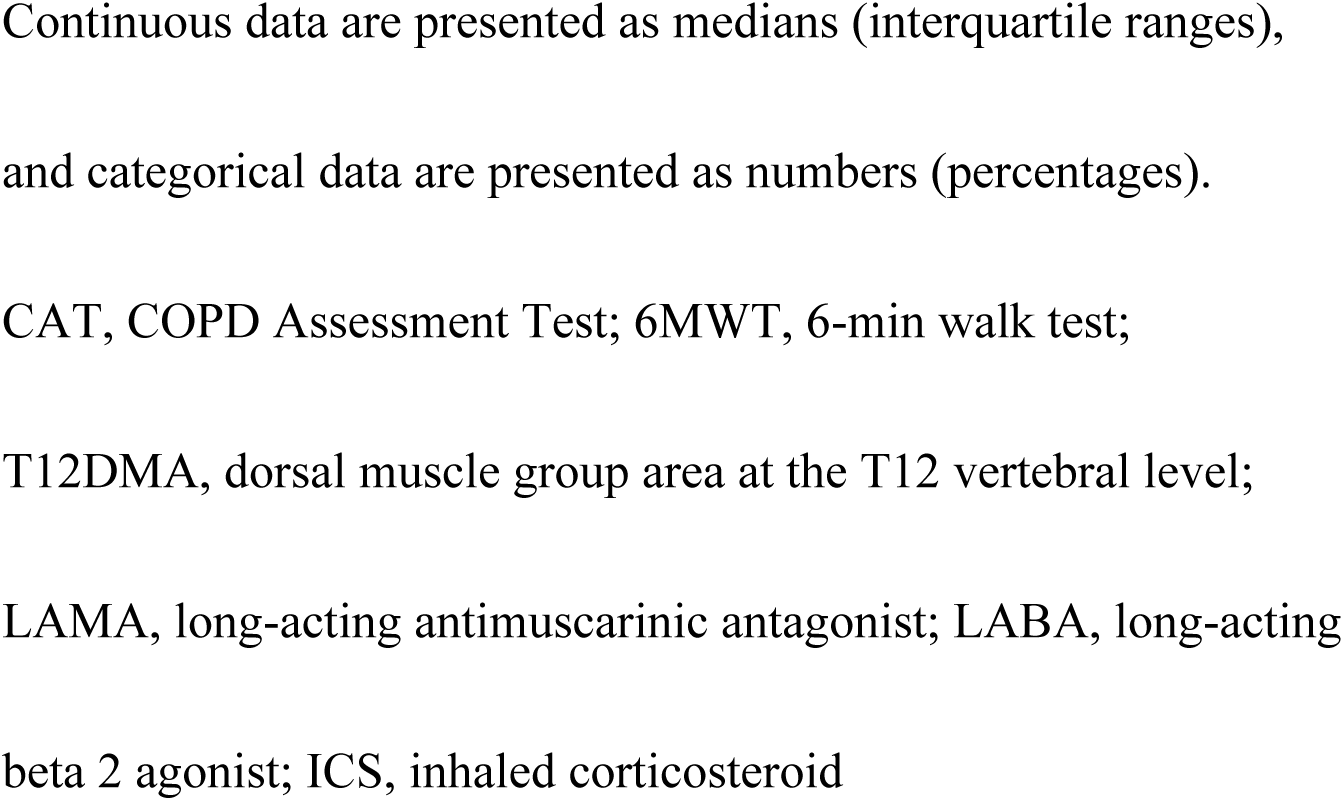
Characteristics of Patients With or Without Sarcopenia Before and After 1:3 Propensity Score Matching.

Given the differences in baseline characteristics between the sarcopenia and non-sarcopenia groups, PSM was performed to reduce the effects of confounding factors. The following variables (age, sex, %FEV1) were included in the PSM model. The numbers of patients in the sarcopenia and non-sarcopenia groups after 1:3 PSM were 62 and 186, respectively, which ensured between-group balance in the distribution of most covariates (Table 2). After PSM, the two groups were well-balanced in terms of baseline characteristics and showed a distribution of characteristics similar to that in the unmatched analysis. As shown in Table 2, the patients in the sarcopenia group demonstrated a shorter walking distance in the 6MWT than those in the non-sarcopenia group (267 m [IQR, 151–343] vs. 345 m [IQR, 247–396], P < 0.001). The median T12DMA indices were 7.53 (IQR, 6.09–8.38) cm^2^/m^2^ and 9.04 (IQR, 7.52–10.30) cm^2^/m^2^ in the sarcopenia and non-sarcopenia groups, respectively (P < 0.001).

As shown in Table 3, the variables included in the multivariate regression logistic models were the T12DMA index (odds ratio [OR] = 0.83; 95% confidence interval [CI], 0.70–0.99, P = 0.042) and BMI (OR = 0.76; 95% CI, 0.68–0.85; P < 0.001). These factors were independent predictors of sarcopenia in patients with COPD.

**Table 3.**
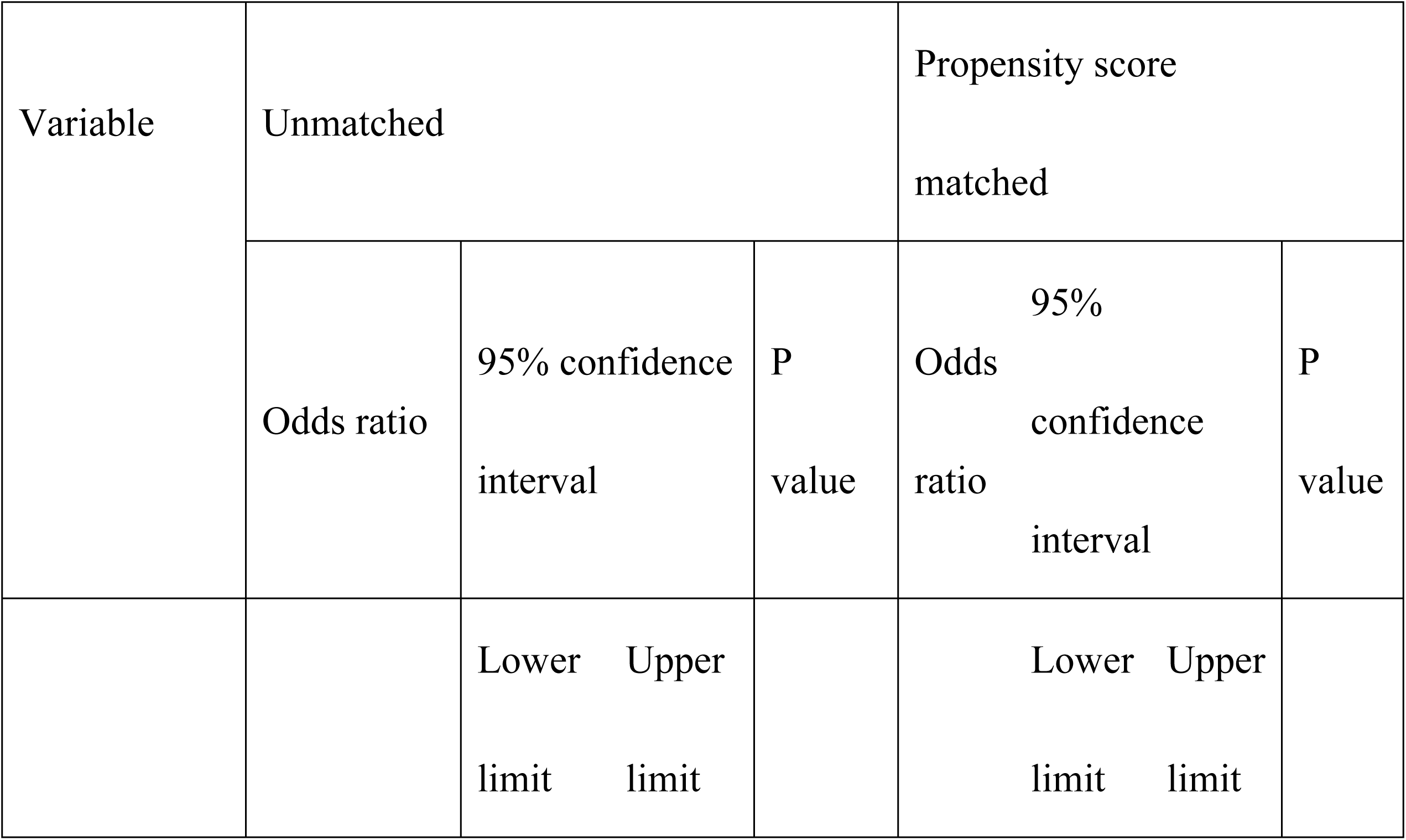
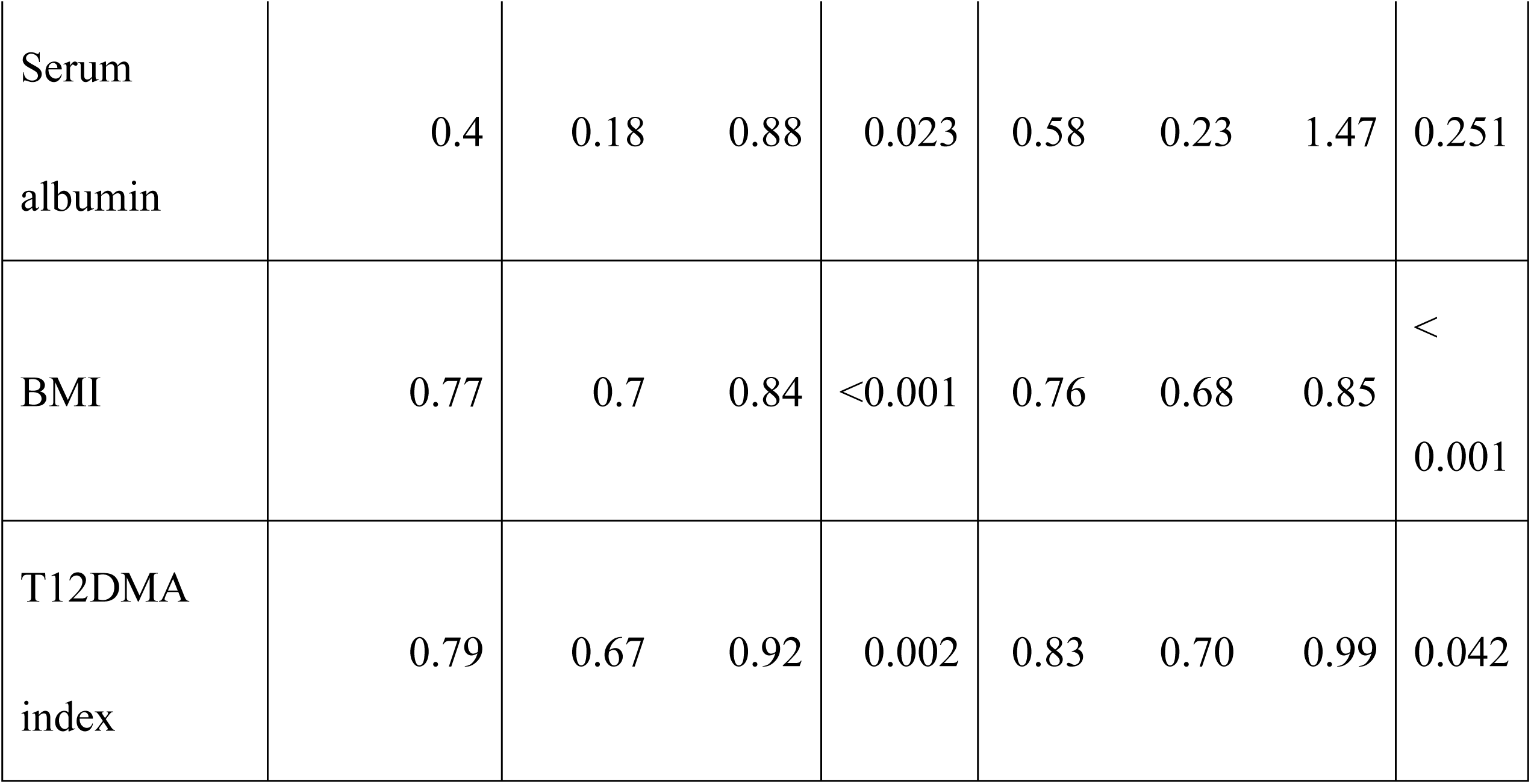
Multivariate Regression Analysis of Associations With Sarcopenia.

## Discussion

In this observational study, we found that the T12DMA index evaluated using chest CT was a predictive factor for sarcopenia in patients with COPD. Furthermore, we establish a reference value for diagnosing sarcopenia using the T12DMA index. The measurement and quantification of sarcopenia using radiological images could be a biomarker for predicting adverse clinical outcomes in patients with COPD.

Sarcopenia is a common comorbidity associated with poor quality of life in patients with COPD [8,9], and the prevalence of sarcopenia in this population is 21.6% [10]. The AWGS 2019 consensus defines sarcopenia as an “age-related loss of SMM, loss of muscle strength, and/or reduced physical performance.” Furthermore, the AWGS 2019 introduced the terms “possible sarcopenia,” defined as low muscle strength with or without reduced physical performance, and “severe sarcopenia” defined by low muscle mass, strength, and physical performance [7]. The diagnosis of sarcopenia is a complex process that requires the specialized evaluation of three components: Muscle mass, strength, and physical function. In 2019, the European Working Group on Sarcopenia in Older People recommended area measurement on a single cross-sectional CT image at the level of the L3 vertebra as an alternative tool to quantify muscles and detect sarcopenia in early stages [11]. Quantification of the L3 skeletal muscle area (SMA) using CT images showed that > 56% of patients with respiratory failure had sarcopenia comorbidities [24].

However, abdominal CT is not a routine examination for patients with COPD, and chest CT, as obtained in regular COPD assessments, does not reach beyond the L1 level. A decline in paravertebral muscle size at T12DMA on chest CT images is associated with mortality in COPD and other diseases [12,13,25]. The DMG area at the T12DMA was used to measure the SMM of patients with COPD to evaluate sarcopenia as an alternative to the L3 SMA. According to studies conducted in healthy individuals, the sarcopenia cut-off values for SMM (SMI) at the T12 level between Asian and US populations differ [14,26]. The discrepancy in these values can be attributed to racial and ethnic differences in lean body mass [27], and the cut-off points vary between studies. Therefore, a consensus on the exact cut-off points that can be used to diagnose sarcopenia is yet to be reached. Furthermore, data regarding the diagnosis of sarcopenia in patients with COPD are lacking. Other studies have also reported that participants with low muscle mass have worse pulmonary function, indicating an important correlation between lung function changes and sarcopenia [28,29]. With aging, respiratory muscle mass declines and function is impaired. A reduction in pulmonary function induces hypoxia, a systemic chronic inflammatory response, and oxidative stress can cause muscle proteolysis and apoptosis, which leads to the loss of SMM [5]. Active screening for sarcopenia, while monitoring lung function, is necessary in elderly patients with COPD. Our study revealed that T12DMA levels decreased and correlated with COPD severity (Fig 3). Patients with COPD with GOLD stage III or IV exhibited significantly lower values than those with stage I. Considering that the decrease in the T12DMA index correlated with the impairment of pulmonary function, we adjusted for potential bias, including predicted %FEV1 by PSM. Our study demonstrated that variables in the multivariate regression logistic models independently predicting sarcopenia in patients with COPD were the T12DMA index (OR = 0.83; 95% CI, 0.69–0.99; P = 0.042) and BMI (OR = 0.76; 95% CI, 0.68–0.85; P < 0.001). Although some studies have reported that low albumin levels are associated with sarcopenia [30,31], we did not observe a significant difference in albumin levels between the sarcopenia and non-sarcopenia groups. Our data offer reference values for the diagnosis of sarcopenia using chest CT scans in Japanese patients with COPD.

The 6MWT is sensitive and reproducible and commonly used to evaluate exercise tolerance [21]. The 6MWT (cut-off < 350 m) predicts mortality and morbidity in patients with COPD and can therefore be used to stratify patients according to their functional capacity [32,33]. Our data and previous reports showed that patients in the sarcopenia group demonstrated a shorter walking distance in the 6MWT than those in the non-sarcopenia group [34]. Physical activity and exercise training are important for maintaining muscle mass and strength in patients with COPD and sarcopenia. A previous study demonstrated that the presence of sarcopenia in patients with COPD did not affect the response to pulmonary rehabilitation and that pulmonary rehabilitation might lead to the resolution of sarcopenia [34]. Therefore, a combination of endurance or interval training and resistance training, which is applied in pulmonary rehabilitation, may be optimal for improving muscle mass, strength, and endurance in patients with sarcopenia and COPD [35,36].

We identified significant and independent correlations between the T12DMA index and sarcopenia. This suggests that SMM analysis using chest CT is a promising indicator for the diagnosis of sarcopenia in patients with COPD. This is because additional instruments, such as a pedometer or a triaxial accelerometer, are not needed to assess physical activity. CT images obtained for other medical purposes can be repurposed to evaluate muscle mass for the diagnosis of sarcopenia, which can be a useful screening tool. The identification of patients with COPD at risk of sarcopenia can contribute to the development of effective preventive and therapeutic measures for sarcopenia-related medical problems.

The primary strengths of our study include its prospective observational design and the inclusion of community-dwelling patients treated by general practitioners in Ishinomaki or surrounding cities, which reflects the real-world COPD population in Japan. However, this study has some limitations. First, the sample size was relatively small. Hence, studies involving larger patient groups are required. Second, a significant proportion of the study population was comprised of men (92%). An insufficient sample size, especially the small number of female patients, may have introduced some bias.

Although this difference reflects the naturally occurring sex distribution among older patients with COPD in Japan, it remains crucial to validate our cut-off values in a larger female cohort.

## Conclusions

Our study offers reference values for the diagnosis of sarcopenia using the T12DMA index evaluated using chest CT in patients with COPD. The identification of patients with COPD in the early stages of sarcopenia can contribute to effective prevention and therapeutic measures, which would lead to better clinical outcomes.

## Data Availability

Data cannot be shared publicly because the dataset contains personally indetifiable information. Ethics Committee (contact via Corresponding author) for researchers who meet the criteria for access to confidential data.

## Acknowledgments

The authors would like to thank the staff of the Outpatient Clinic of the Japanese Red Cross Ishinomaki Hospital, Ishinomaki, Japan, for their help with data management. We are also grateful to the healthcare professionals affiliated with the ICON for their kind help and cooperation. We are also grateful to the healthcare professionals affiliated with the ICON for their kind help and cooperation with this study. We would also like to thank Editage (www.editage.jp) for English language editing.

## Notes

### Competing Interest Statement

The authors have declared no competing interest.

### Funding Statement

The author(s) received no specific funding for this work.

### Author Declarations

The Ethics Committee of the Japanese Red Cross Ishinomaki Hospital

